# Impact of opioid agonists on mental health in substitution treatment for opioid use disorder: A systematic review and Bayesian network meta-analysis of randomized clinical trials

**DOI:** 10.1101/2020.07.04.20146506

**Authors:** Ehsan Moazen-Zadeh, Kimia Ziafat, Kiana Yazdani, Mostafa Mamdouh, James Wong, Amirhossein Modabbernia, Peter Blanken, Uwe Verthein, Christian G. Schütz, Kerry Jang, Shahin Akhondzadeh, R. Michael Krausz

## Abstract

**Objective:** There is a dearth of high-quality systematic evidence on the impact of opioid substitution medications on mental health. We compared mental health outcomes between opioid medications and placebo/waitlist, and between different opioids.

**Methods:** This systematic review and meta-analysis of randomized clinical trials (RCTs) was pre-registered at PROSPERO (CRD42018109375). Embase, MEDLINE, PsychInfo, CINAHL Complete, and Web of Science Core Collection were searched from inception to May 2020. RCTs were included if they compared opioid agonists with each other or with a placebo/waitlist in substitution treatment of patients with opioid use disorder, and reported at least one mental health outcome on a span of more than 1-month post baseline. Studies with psychiatric care, adjunct psychotropic medications, or unbalanced psychosocial services were excluded. Primary outcomes were comparison of depressive symptoms and overall mental health between opioids and placebo/waitlist. Random effects model was used for all the meta-analysis.

**Results:** Nineteen studies were included in the narrative synthesis and 15 in the quantitative synthesis. Hydromorphone, diacetylmorphine (DAM), methadone, slow-release oral morphine, buprenorphine, and placebo/waitlist were among the included interventions. Based on network meta-analysis for primary outcomes, buprenorphine (SMD (CI95%)= −0.61 (−1.20, −0.11)), DAM (−1.40 (−2.70, −0.23)), and methadone (−1.20 (−2.30, −0.11)) were superior to waitlist/placebo on overall mental health. Further direct pairwise meta-analysis indicated that overall mental health improved more in DAM compared to methadone (−0.23 (−0.34, −0.13)).

**Conclusions:** It appears that opioid medications improve mental health independent of psychosocial services. Potential contribution of other factors needs to be further investigated.

## Introduction

In recent years, there has been a significant increase in illicit opioid use and opioid-related fatalities worldwide with illicit opioids accounting for the greatest harm to the health of users among all other substances of abuse (1). In the US, 64.3% of adults with opioid use disorder suffer from a current comorbid mental illness (2). Rates of comorbid mental disorders among patients seeking opioid substitution treatment are 20-80% in Europe and approximately 80% in Ontario, Canada (3,4). Despite of these high rates of comorbidity, only around 1/4 of adults with concurrent opioid use disorder and acute mental illness receive treatment for both problems (2). Furthermore, the array of psychosocial interventions studied in treatment of opioid use disorder have produced mixed results in terms of improving mental health status (3,5). The results of using anti-depressants for treatment of patients with comorbid major depression opioid use disorder are inconsistent (3).

Understanding the dynamics of potential factors that contribute to the improvement of mental health in patients with opioid use disorder is essential for the development of novel effective interventions. Opioid agonist medications are the mainstay and first line of treatments in opioid use disorder and have a long history in the treatment of mood disorders even before anti-depressants became available on the market (6,7). Recent clinical studies demonstrated the efficacy of buprenorphine in improving depression and risk of suicide in patients with refractory major depression (8), and several lines of evidence suggest involvement of the endogenous opioid system in mood and anxiety disorders (9,10). Based on our literature search, there have been only two systematic reviews on mental health-related outcomes in opioid substitution treatment (11,12). While both studies reported improved mental health outcomes in opioid substitution treatment, the scope their review was limited and they were unable to distinguish between effects of opioid agonists on mental health from the potential effects of adjunctive psychosocial interventions because they lacked any comparison between treatment arms.

In order to obtain greater clarity of the potential role of opioid medications in improving mental health in patients with opioid use disorder, we conducted a comparative assessment of mental health outcomes between opioid agonist medications and control conditions (i.e. placebo or waitlist), in clinical trials of opioid substitution treatment. The aggregated results of direct comparisons of mental health outcomes between different opioid agonists are also provided.

## Materials and methods

### Protocol

The PRISMA guidelines and the latest version of Cochrane Handbook were employed throughout the whole conduct and reporting of the study (13). All methods (https://www.crd.york.ac.uk/prospero/, CRD42018109375) were predefined and registered before initiation of the screening phase.

### Research question and inclusion/exclusion criteria

The primary question was whether mental health outcomes improve in active substitution treatment with opioid agonists more than neutral control conditions such as placebo/waitlist independent of psychosocial interventions in patients with opioid use disorder. The secondary question was that whether mental health outcomes differentially improve during treatment with diverse opioid agonists in patients with opioid use disorder. Randomized clinical trials were included if they compared any opioid agonists with each other or with a placebo/waitlist in substitution treatment of patients with opioid use disorder and reported at least one mental health outcome using a validated measurement tool on a span of more than 1 month post-baseline. Studies were excluded if patients received comprehensive psychiatric care or were randomized to adjunctive interventions such as focused psychotherapy or adjunctive psychotropic medications. Ancillary routine counselling was acceptable if it was available to all the participants in a study and was provided in the same way across all treatment arms.

### Search strategy

The following general combination of search terms and Boolean operators were used: [names of all known opioid substitution medications separated by OR] AND [terms related to clinical trials separated by OR] AND [terms related to mental health separated by OR]. The exact search strategy used to search each database is presented in Supplementary Table 1. On September 10, 2018, a comprehensive list of databases were searched including: EBM Reviews - Cochrane Central Register of Controlled Trials, Embase, MEDLINE(R) and Epub Ahead of Print, In-Process & Other Non-Indexed Citations and Daily, PsychInfo, CINAHL Complete, Web of Science Core Collection, LILACS, OpenGrey, Google Scholar first 200 citations, clinicaltrials.gov and clinicaltrialsregister.eu for the completed/terminated trials registered in the recent 5 years. Finally, a hand search of reference lists from included trials as well as major systematic reviews of substitution treatments was performed to find additional full-texts. Experts in the field were also consulted. In May 2019 and May 2020, we updated our search to find any relevant newly published studies by searching the literature post September 2018 and asking experts in the field.

**Table 1.**
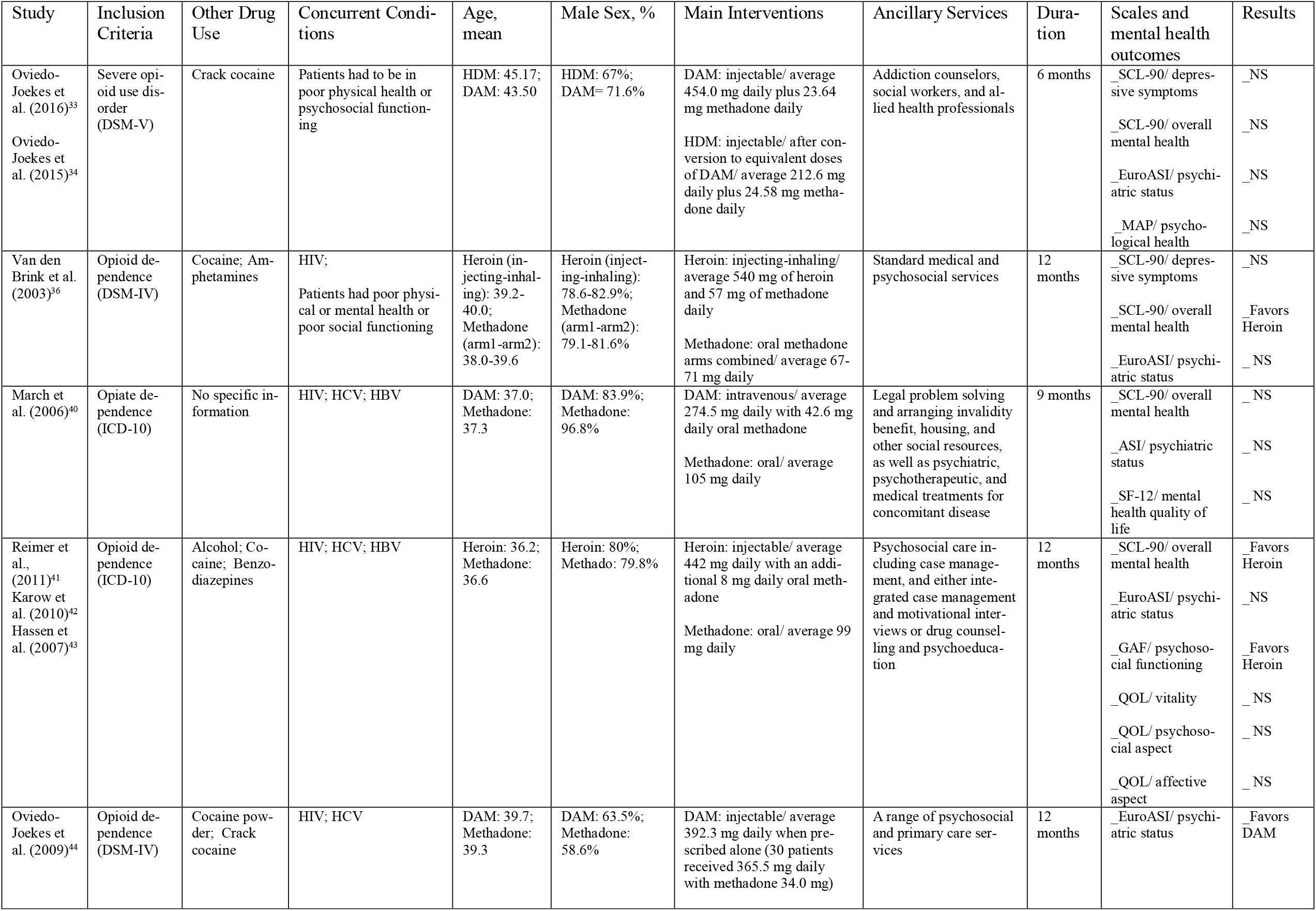
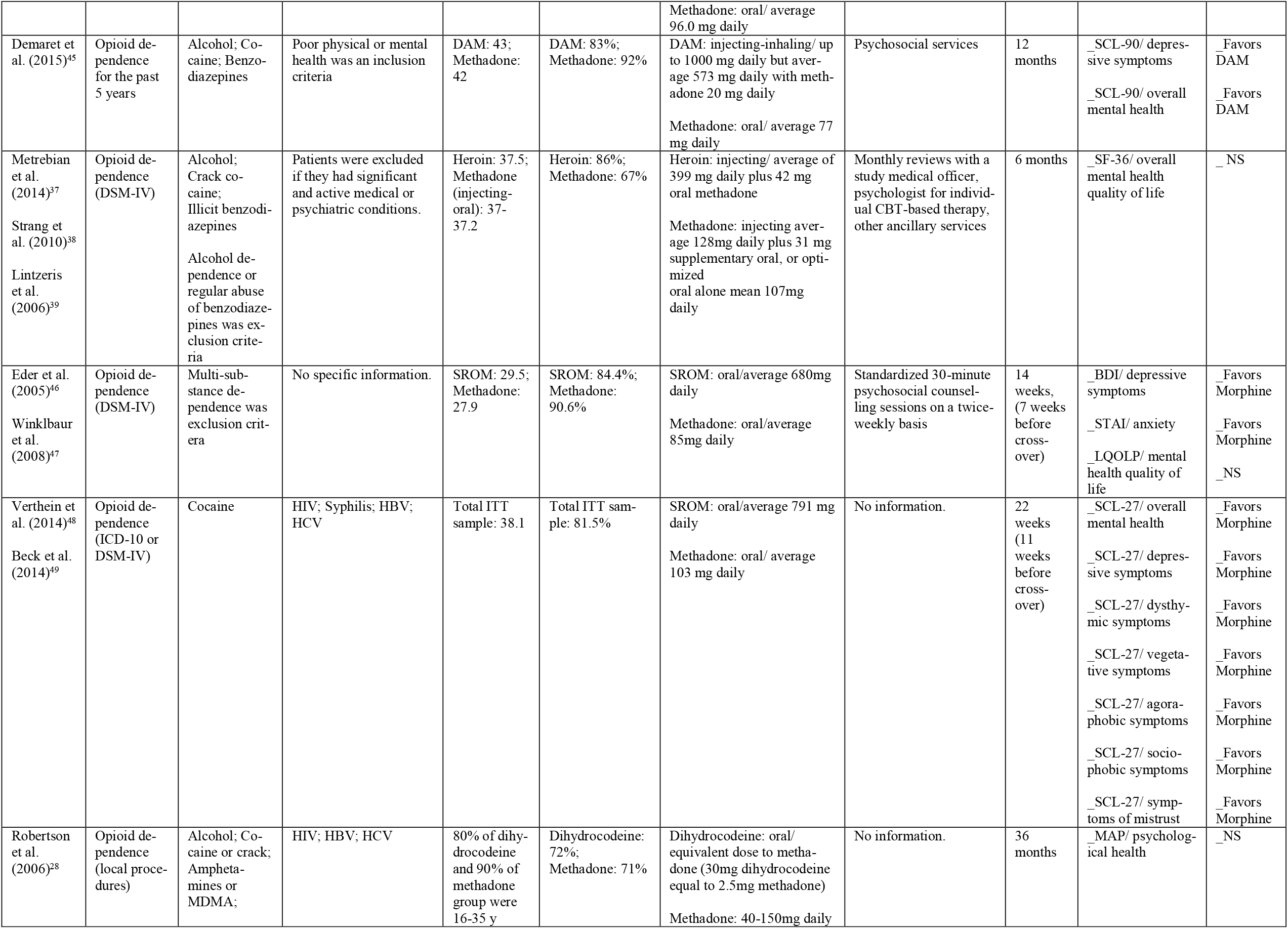
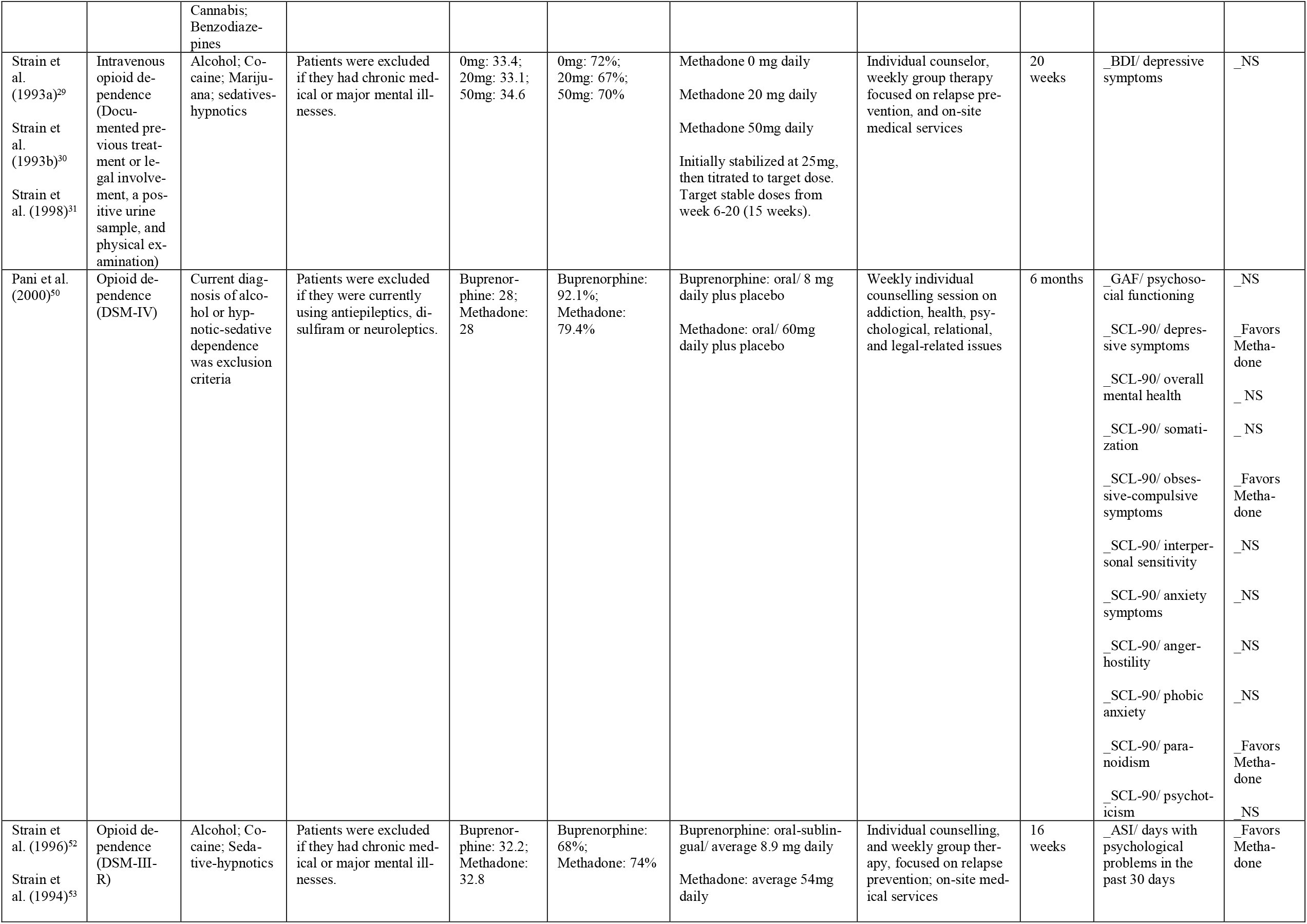
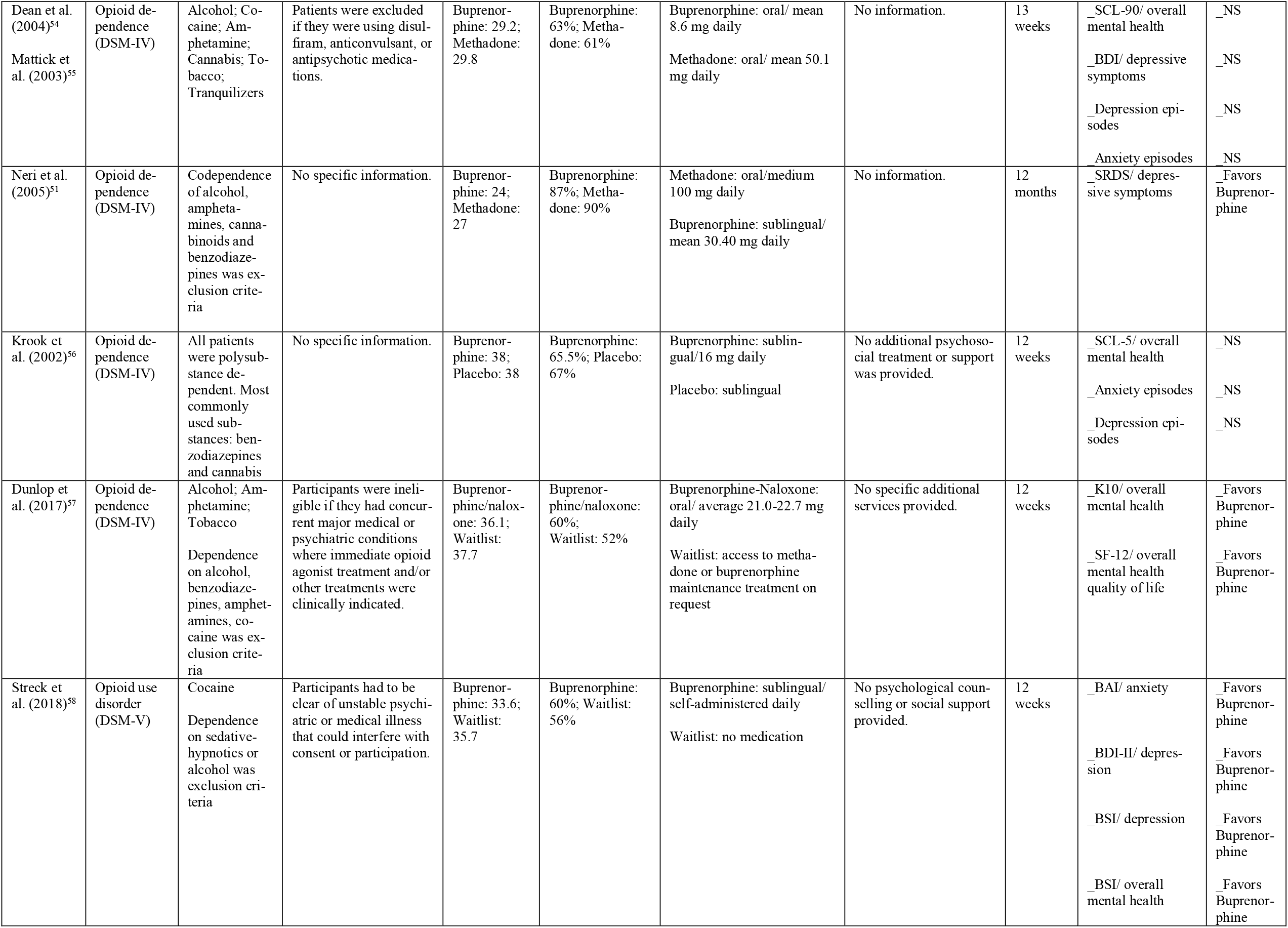
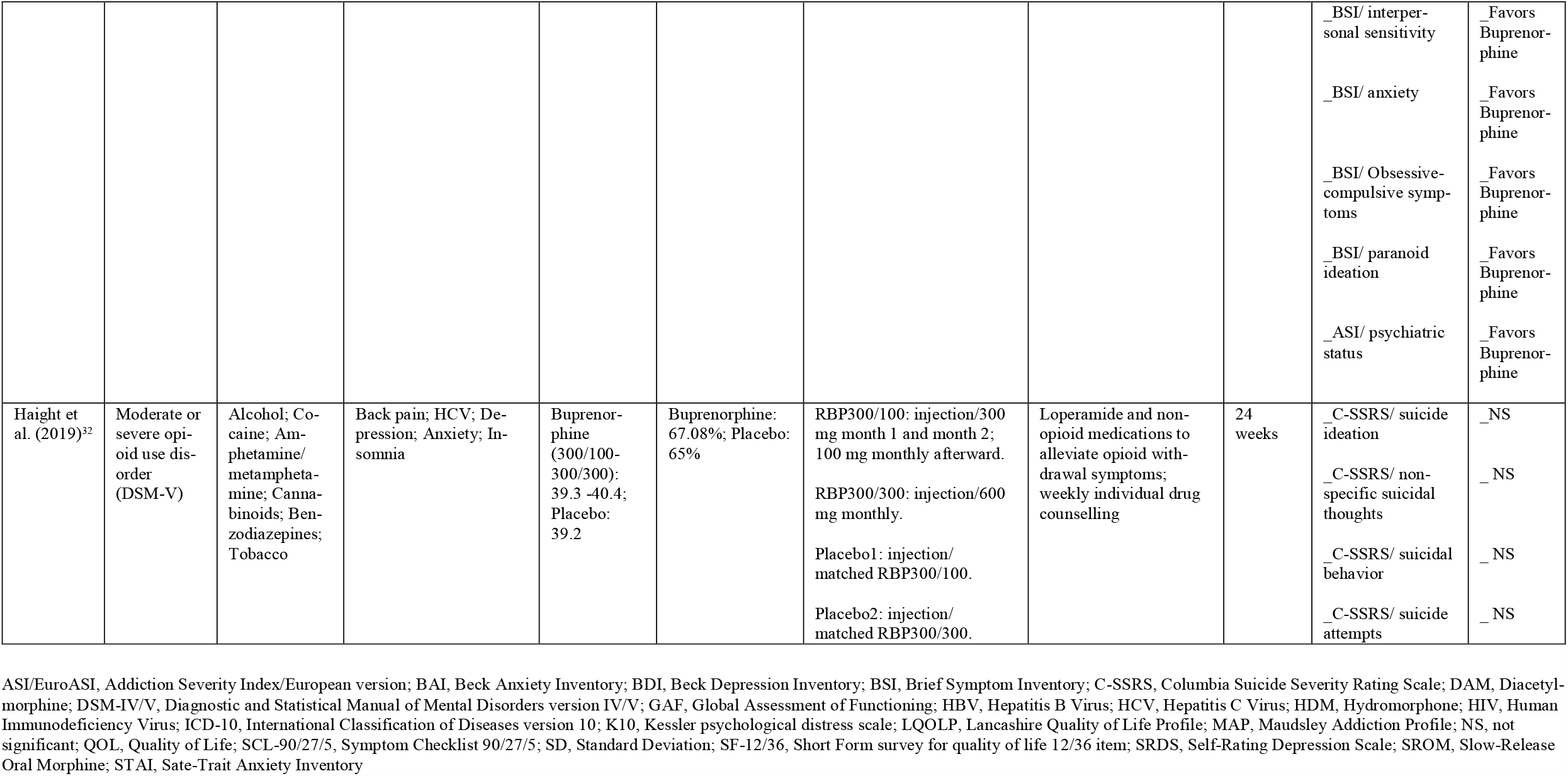
Summary of studies included in the systematic review of mental health outcomes in opioid substitution treatment

### Study selection

Four authors worked in parallel, two by two, to screen all the retrieved citations after they were calibrated for the specific inclusion/exclusion criteria using a sample of 20 challenging citations selected by the lead investigator and the senior investigator. These citations were assessed by all four authors and discussed with the lead investigator and senior investigator to reach a consensus regarding any disparities in assessment results. Then, the retrieved citations from the systematic search stage were equally distributed in half among the two pairs of authors. In a two stage process, authors first screened the titles/abstracts, and then screened the full-text of citations. At each stage, discrepancies in the results were resolved between the screeners after reaching a consensus with both the lead investigator and senior investigator. Conference abstracts, thesis reports, and registries of clinical trials were also included if enough data could be collected, either through the reports themselves or by contacting the authors.

### Data extraction

Four authors carried out data extraction on a primary sample of 4 studies, where results were compared, discussed, and calibrated amongst them and the lead investigator. Afterwards, the 4 authors worked in parallel pairs to extract data from all studies. Final results were compared, and in cases of discrepancy, a consensus was reached after discussion with the lead investigator and senior investigator. Relevant data were extracted from the included studies using excel sheets with predefined columns including those mentioned in Table 1 as well as details of analytical methods and reported statistics. For each mental health outcome, we recorded: primary or secondary, scale of measurement, baseline and follow-up values, time-line of measurements, and any summary measures reported plus statistical tests that were used.

### Risk of bias assessment

Four authors conducted a risk of bias assessment on a primary sample of 5 studies in which results were compared, discussed, and calibrated amongst them and the lead investigator. This was followed by the two pairs of authors working in parallel and assessing the risk of bias in all studies. Final results were compared, and in cases of discrepancy, a consensus was reached after discussion with the lead investigator and senior investigator. The recently released comprehensive Cochtrane Risk of Bias Tool version 2 (RoB2), which evaluates each outcome for 6 domains of potential bias as well overall risk of bias, was used for assessment of single studies (14). In cases of similarity among the assessment results for the different outcomes of a single study, a single assessment result was reported for the whole study.

### Outcomes, synthesis strategy, and measures

Primary outcomes included standardized mean difference in score changes from baseline to endpoint between opioid agonists and placebo/waitlist for depressive symptoms and overall mental health symptomatology. Secondary outcomes included standardized mean difference in score changes among different opioid agonists for any measures of mental health. Narrative synthesis was carried out by putting together studies on direct comparison of each specific pair of medications, e.g. methadone and buprenorphine. Characteristics of those studies as well as the reported mental health outcomes were used for this synthesis. Quantitative synthesis was carried out in two stages; assessing primary outcomes using network metaanalysis, and direct pairwise meta-analysis for secondary outcomes.

For the quantitative synthesis only data from the following measures of mental health were combined: depression subscale of Symptom Checklist-90 (SCL-90) as well as shorter versions, Beck Depression Inventory (BDI), or Self Rating Depression Scale (15,16); total scores/global severity index from SCL-90 as well as shorter versions, Brief Symptom Inventory (BSI), or Kessler psychological distress scale (K10) (17-19); composite scores of psychiatric status on Addiction Severity Index (ASI) or European ASI (20); and mental health quality of life measured by Short Form Health Survey-36 (SF36) or Lancashire Quality of Life Profile (LQOLP) (21,22). Validity, reliability, and level of correlation of similar measures are discussed elsewhere (15-22).

### Statistical analysis

All the analyses were pre-planned. Data from injecting, inhaling, and oral administration of medications or from different doses of a single medication were combined for the studies that included more than one administration route or dosing.

For the network meta-analysis (NMA), standardized mean differences (SMD) and standard errors (SE) that were estimated for each single study on each included mental health outcome were used as the input. An online free version of R package GeMTC for Bayesian NMA (https://gemtc.drugis.org/) by Markov Chain Monte Carlo was used with all the codes accessible from the website. Contrast-based NMA was conducted. Random effects method was used rather than fixed effects in order to account for the heterogeneity among studies and potential inconsistencies throughout the network; however, results from both methods were presented. Normal likelihood and identity link function were assumed. Number of chains were set at 4, burn-in iterations at 5000, inference iterations at 20000, and thinning factor at 10. Convergence diagnostics were used to check for the effect of starting values and proper mixing of chains. Leverage-residual deviance plots were used to decide if the final model was appropriate and measures of model fit such as Deviance Information Criterion (DIC) were reported. Node-splitting analysis of consistency was not applicable as there was no closed loop in the model network. Rank probabilities plot and effect estimates were used to compare the medications with placebo/waitlist. The quality of evidence for the network meta-analysis was also assessed using guidelines defined by Salanti et al. (23), based on the GRADE guidelines for assessment of quality of evidence and overall rating of confidence in the estimates (24).

For the direct pairwise meta-analysis, pooled estimates of SMDs were provided alongside SEs. Comprehensive Meta-analysis v2.0 was utilized for this purpose, using random effects model. Measures of heterogeneity were reported including Q statistic for assessing the significance of heterogeneity, the I^2^ to evaluate the proportion of total variability attributable to heterogeneity, and Tau^2^ for the extent of heterogeneity. Funnel plots were presented if there were enough studies to make the assessment of small study effects, including publication bias, feasible. Symmetry of funnel plots does not eliminate possibility of publication bias, but asymmetry of plots warrants further scrutiny. By definition, publication bias refers to a systematic bias in the probability of studies getting published based on the significance and direction of their results. Sensitivity analysis was considered if there were more than one study with low risk of bias on a specific outcome. Pooled estimates for these low risk studies would be compared with the combination of low-risk/high-risk studies.

## Results

### Study Selection and overview

After duplicate removal, 2983 citations were screened (Figure 1). Twenty-two studies initially met inclusion criteria, but only 19 were included in this review because sufficient data were not received from authors of three studies (25-27). Furthermore, only 15 studies were included in quantitative synthesis as outcome from two studies were not reported with adequate details (28-31), and similar measures to the ones used in studies by Haight et al. (32), and Strain et al. (52,53), were not available from other studies in order to be included for quantitative synthesis.

**Fig. 1.**
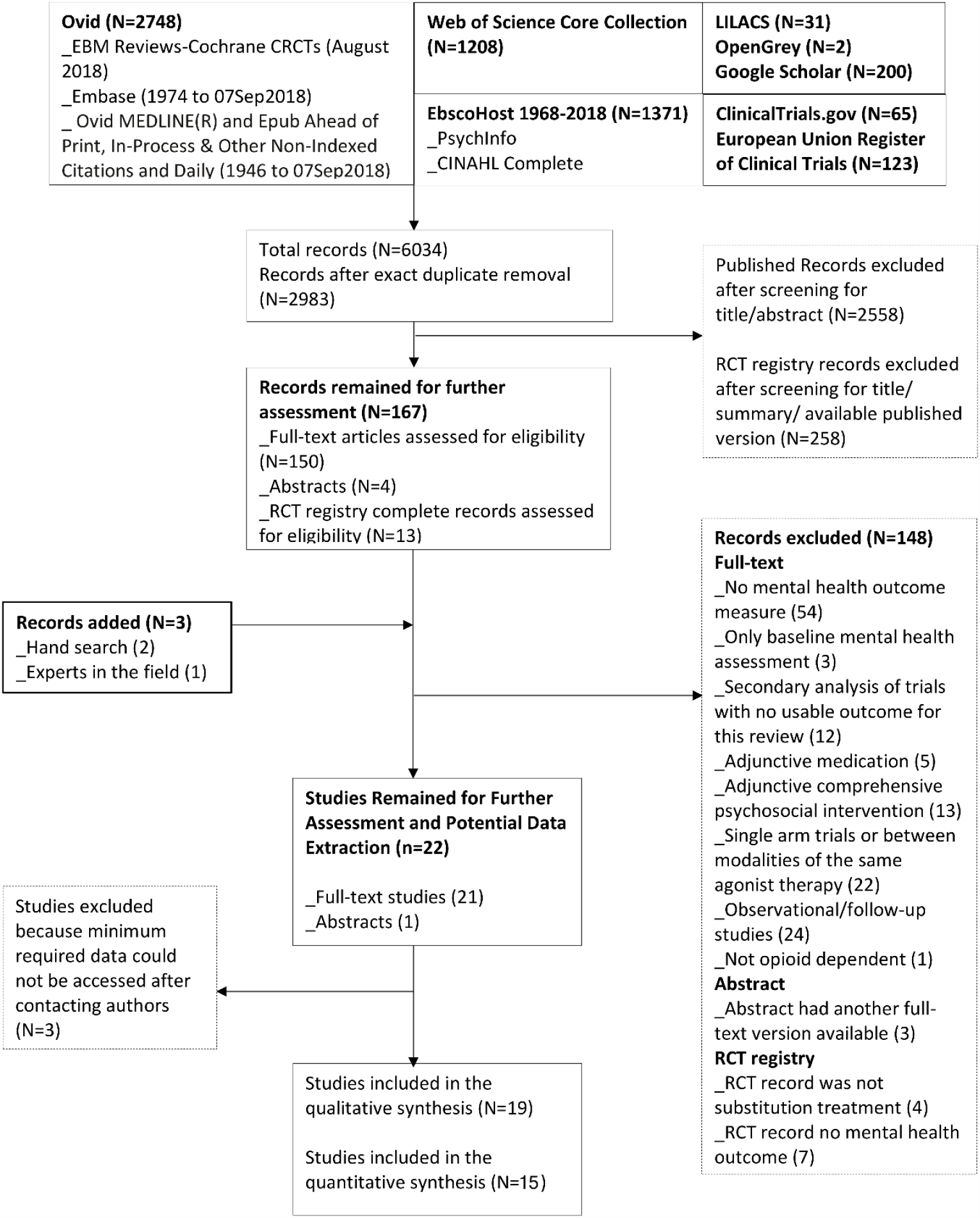
PRISMA flow diagram of studies assessed for systematic review of clinical trials of opioid substitution treatment.

Table 1 represents study characteristics and any mental health outcome reported in 19 included studies. Table 2 represents risk of bias assessment for the 19 included studies. The results of meta-analysis are reported in Table 3 as well as Figures 2-5.

**Table 2.**
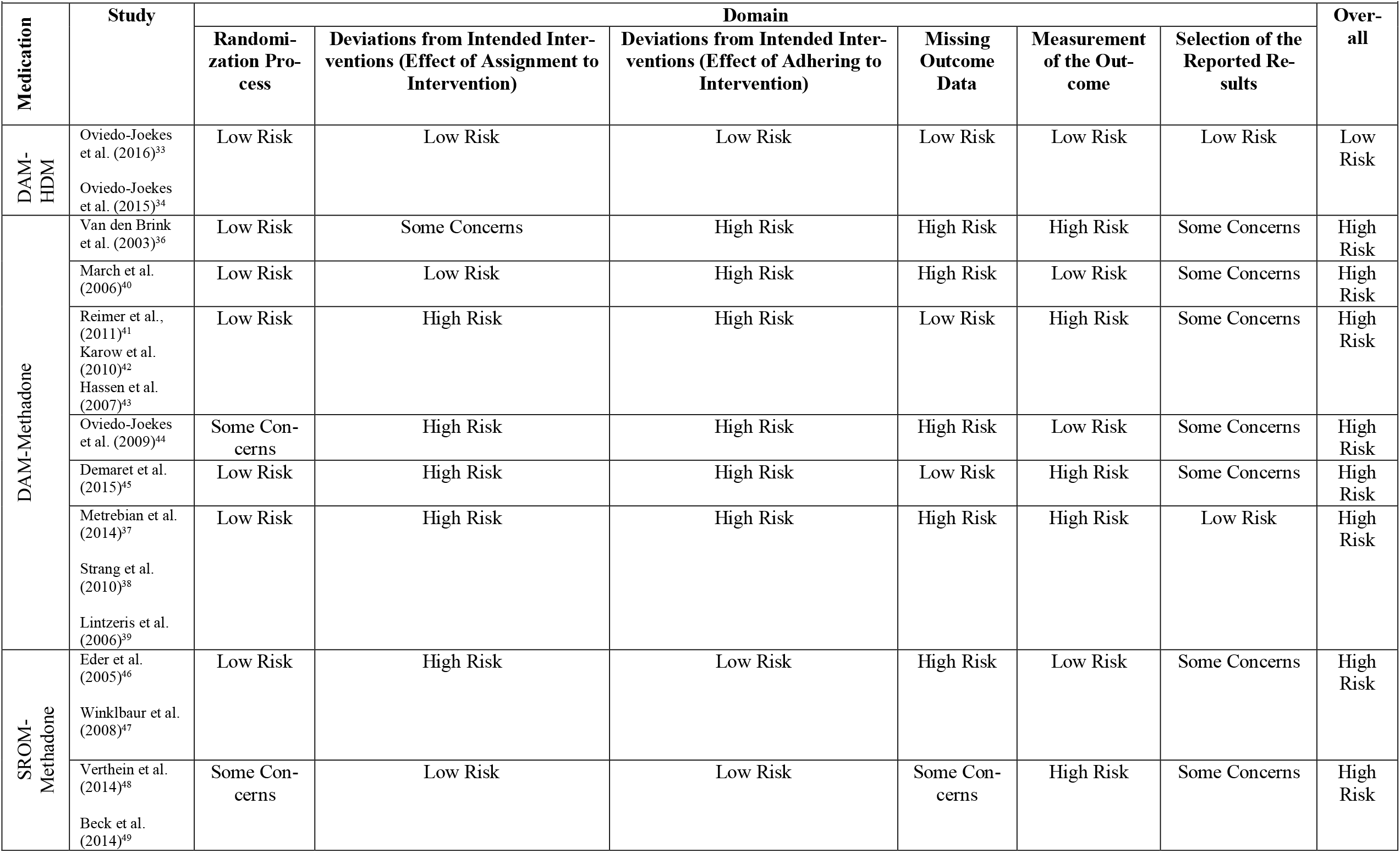
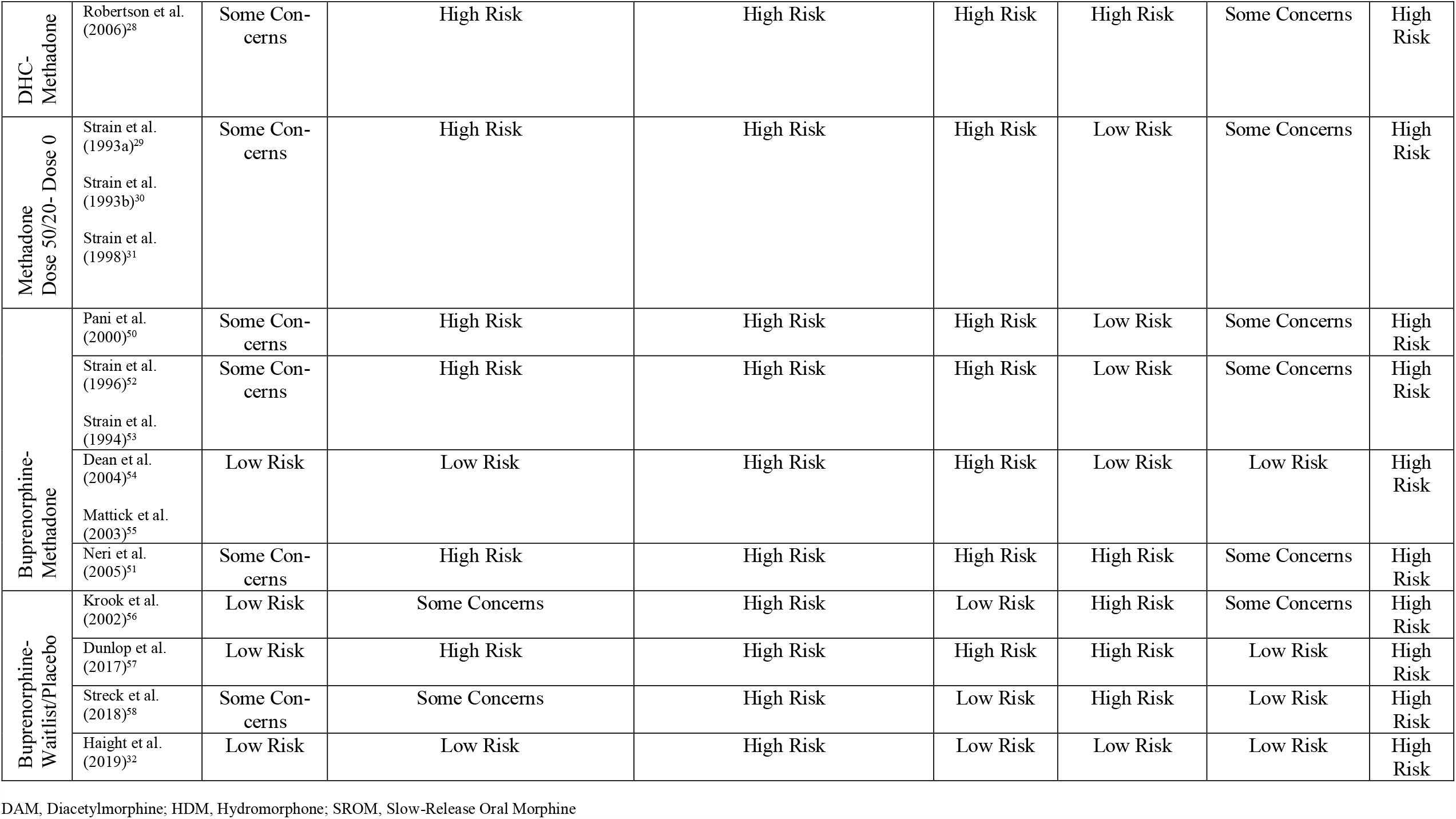
Risk of Bias Assessment for mental health outcomes in clinical trials of opioid substitution treatment

**Table 3.**
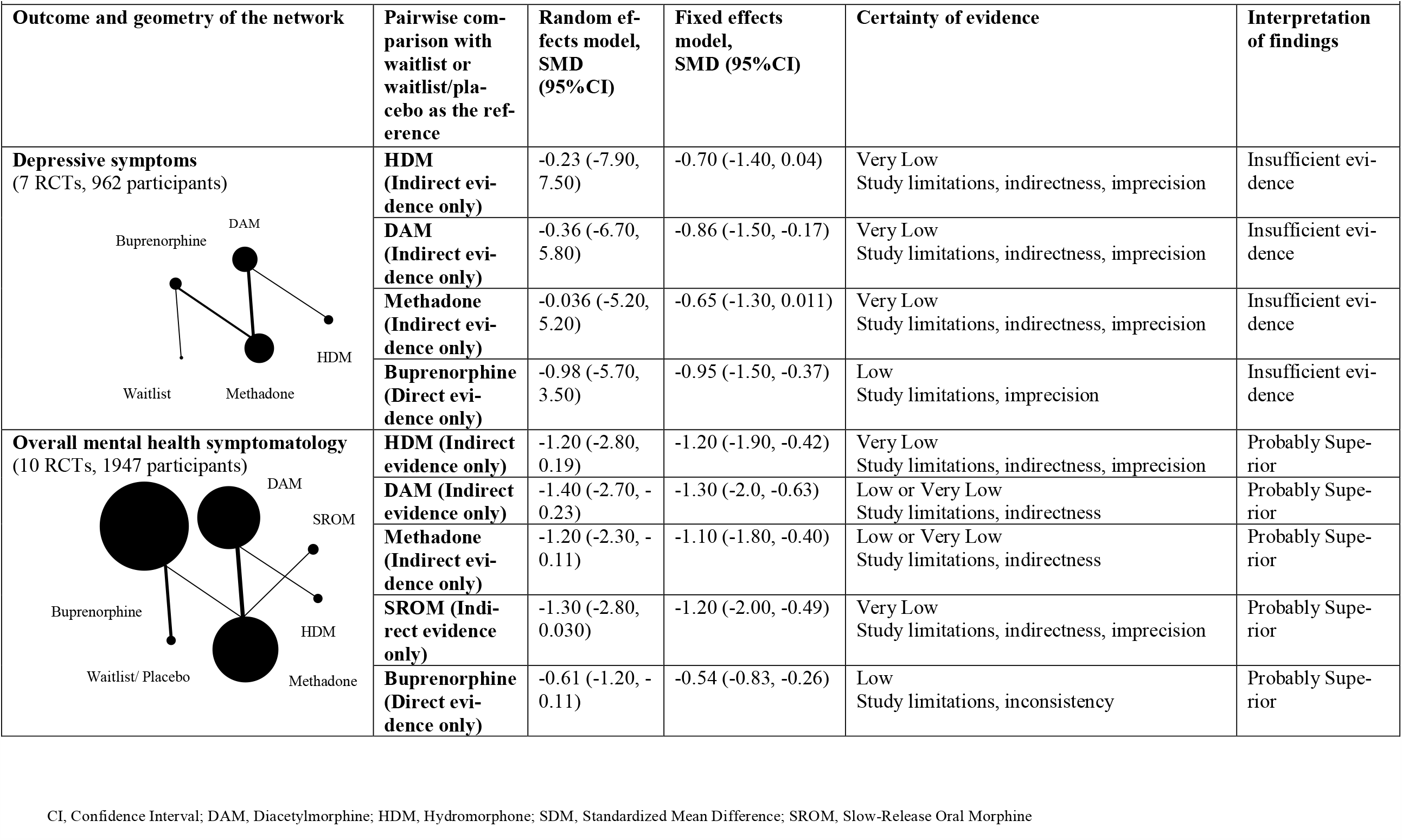
Summary of findings from network meta-analysis and GRADE assessment for mental health outcomes in opioid substitution treatment

|  |  |  |  |  |  |
| --- | --- | --- | --- | --- | --- |
| Outcome and geometry of the network | Pairwise comparison with waitlist or waitlist/placebo as the reference | Random effects model, SMD (95% CI) | Fixed effects model, SMD (95% CI) | Certainty of evidence | Interpretation of findings |
| **Depressive symptoms**<br>(7 RCTs, 962 participants) | **HDM (Indirect evidence only)** | -0.23 (-7.90, 7.50) | -0.70 (-1.40, 0.04) | Very Low<br>Study limitations, indirectness, imprecision | Insufficient evidence |
|  | **DAM (Indirect evidence only)** | -0.36 (-6.70, 5.80) | -0.86 (-1.50, -0.17) | Very Low<br>Study limitations, indirectness, imprecision | Insufficient evidence |
|  | **Methadone (Indirect evidence only)** | -0.036 (-5.20, 5.20) | -0.65 (-1.30, 0.011) | Very Low<br>Study limitations, indirectness, imprecision | Insufficient evidence |
|  | **Buprenorphine (Direct evidence only)** | -0.98 (-5.70, 3.50) | -0.95 (-1.50, -0.37) | Low<br>Study limitations, imprecision | Insufficient evidence |
| **Overall mental health symptomatology**<br>(10 RCTs, 1947 participants) | **HDM (Indirect evidence only)** | -1.20 (-2.80, 0.19) | -1.20 (-1.90, -0.42) | Very Low<br>Study limitations, indirectness, imprecision | Probably Superior |
|  | **DAM (Indirect evidence only)** | -1.40 (-2.70, -0.23) | -1.30 (-2.0, -0.63) | Low or Very Low<br>Study limitations, indirectness | Probably Superior |
|  | **Methadone (Indirect evidence only)** | -1.20 (-2.30, -0.11) | -1.10 (-1.80, -0.40) | Low or Very Low<br>Study limitations, indirectness | Probably Superior |
|  | **SROM (Indirect evidence only)** | -1.30 (-2.80, 0.030) | -1.20 (-2.00, -0.49) | Very Low<br>Study limitations, indirectness, imprecision | Probably Superior |
|  | **Buprenorphine (Direct evidence only)** | -0.61 (-1.20, -0.11) | -0.54 (-0.83, -0.26) | Low<br>Study limitations, inconsistency | Probably Superior |

CI, Confidence Interval; DAM, Diacetylmorphine; HDM, Hydromorphone; SDM, Standardized Mean Difference; SROM, Slow-Release Oral Morphine

**Fig. 2.**
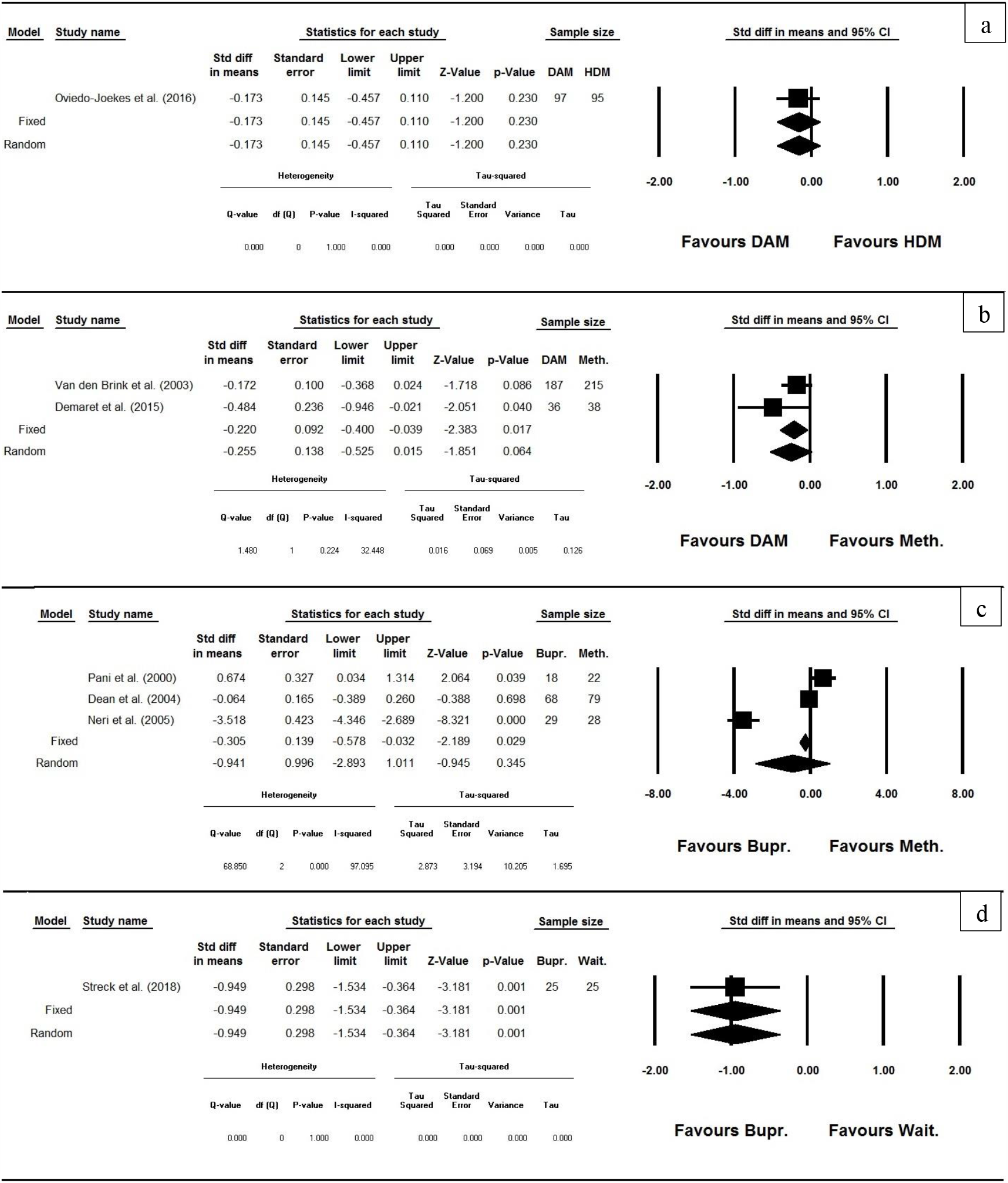
Meta-analysis of depressive symptoms in clinical trials of opioid substitution treatment.

**Fig. 3.**
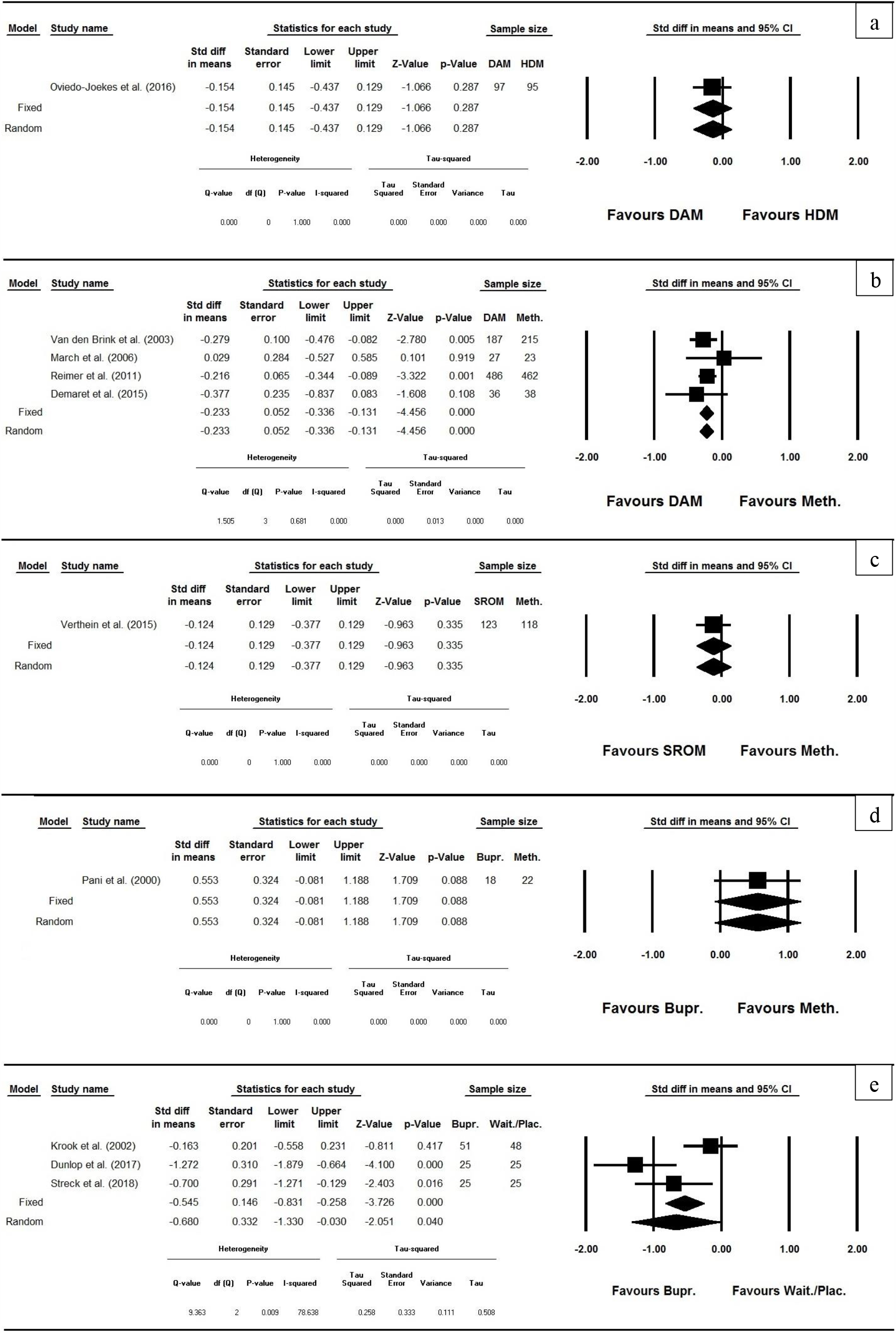
Meta-analysis of overall mental health symptomatology in clinical trials of opioid substitution treatment.

**Fig. 4.**
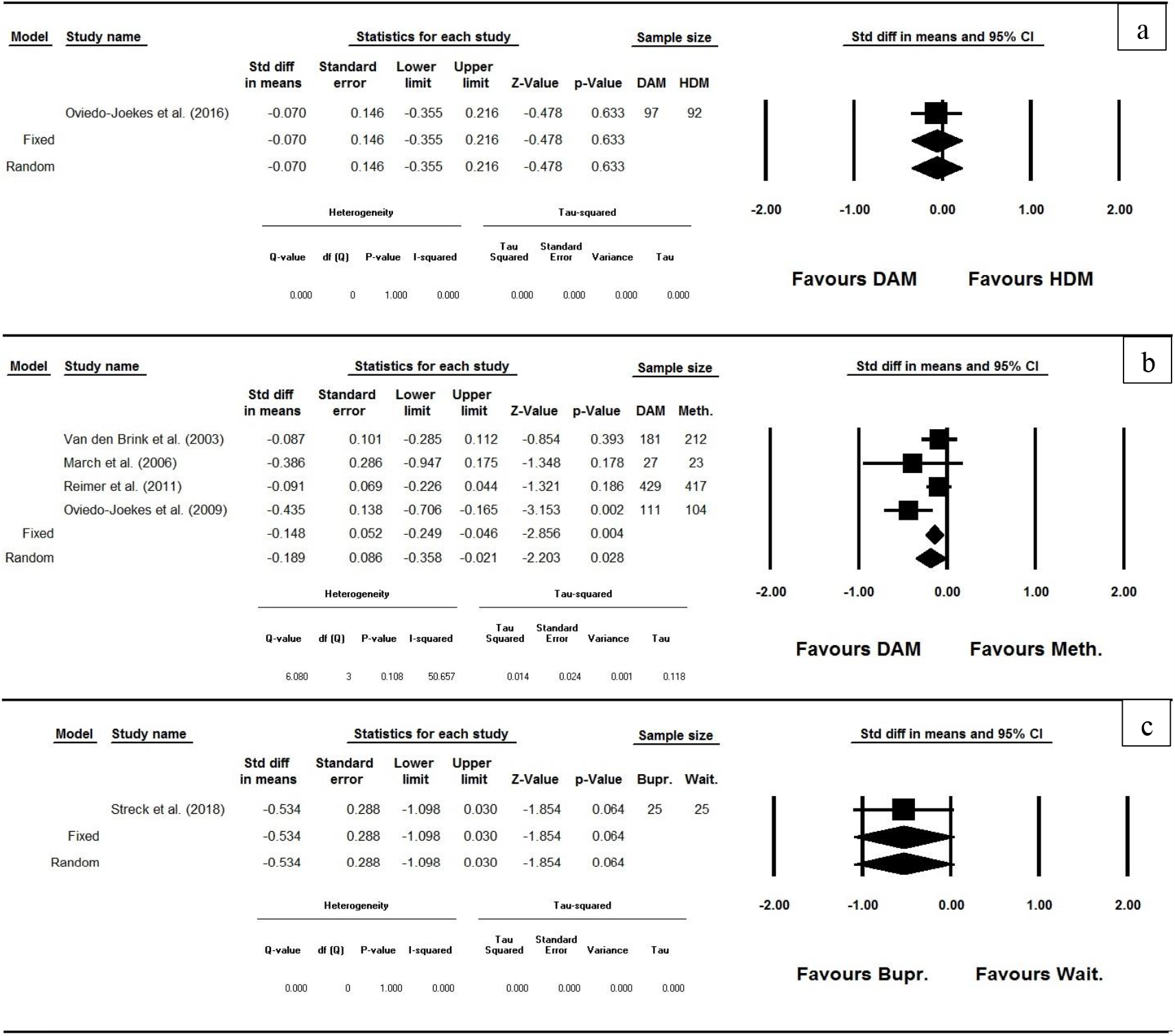
Meta-analysis of addiction severity index in clinical trials of opioid substitution treatment.

**Fig. 5.**
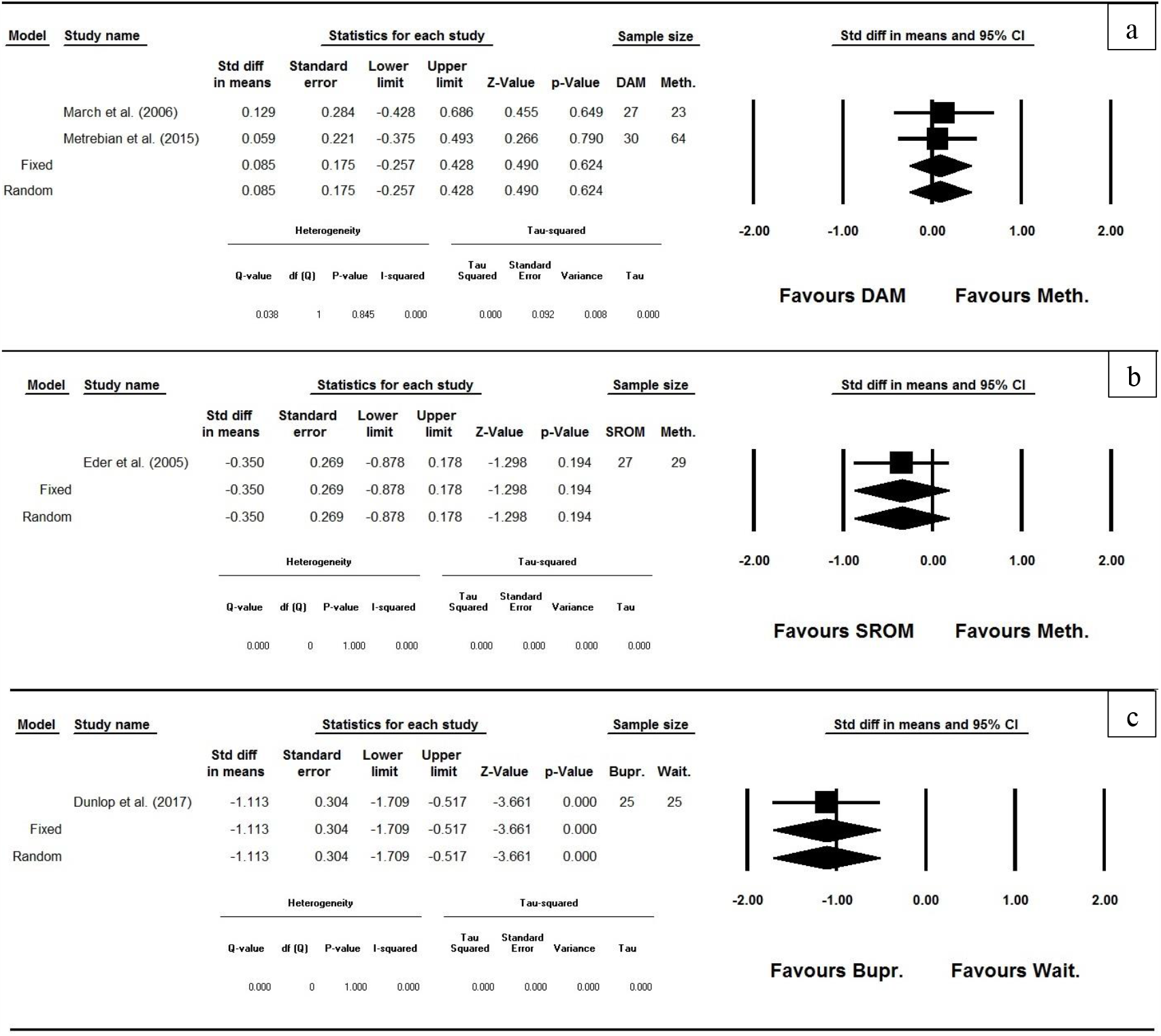
Meta-analysis of mental health quality of life in clinical trials of opioid substitution treatment.

### Risk of Bias

Except for one study with Low Risk of bias in all the RoB2 indicated domains (33,34), all studies had Some Concerns or High Risk of bias in multiple assessment domains, which resulted in an overall High Risk of bias for those studies (Table 2). The Randomization Process domain had the highest frequency of Low Risk studies, while the two domains concerning Deviations from Intended Interventions had the highest frequency of High Risk studies (Table 2). In this review, small study effects was not assessed due to the low number of studies in each pairwise comparison according to Cochrane’s recommendation of the availability of around 10 trials on each pairwise comparison for a meaningful interpretation of small study effects in particular publication bias (13,35).

### Narrative synthesis

#### Hydromorphone-Diacetylmorphine

Only one study was included comparing injectable HDM with injectable DAM in patients with severe opioid use disorder and poor physical/psychosocial conditions, most of whom had a history of relapse after previous methadone treatments (Table 1) (33,34). Oral methadone and counselling services were available to all participants. No significant difference was reported in improvement of mental health outcomes, i.e. depressive symptoms and overall mental health changes were similar between the two treatments after 6 months of therapy (Table 1).

#### Diacetylmorphine-Methadone

Six studies compared injectable heroin (DAM) with oral methadone, while in 2 of those studies inhalable heroin and injectable methadone were provided as well (Table 1) (36-39). In all studies patients were provided with psychosocial care, and those in the heroin treatment had access to optional oral methadone. Most patients had a history of relapse after prior methadone treatments and were polydrug users. Patients in 4 studies had poor physical/psychosocial health conditions or high levels of serious comorbid medical conditions like HIV and HCV (36,40-44). Duration of studies varied from 6 months to 12 months. While 2 studies mainly reported no significant differences between the two treatments on various mental health scales (36,37,40,41), 4 studies reported superiority of DAM on some scales, particularly overall mental health (Table 1) (36,41,44,45).

#### Dihydrocodeine-Methadone

Only one study was included comparing oral dihydrocodeine and oral methadone in patients with opioid dependence (Table 1) (28). Majority of patients were using other comorbid substances at baseline. Prevalence of serious comorbid conditions was low in the sample. No significant difference was reported in the improvement of psychological health after 36 months of therapy (Table 1).

#### Slow-release oral morphine (SROM) -Methadone

Two studies compared SROM with oral methadone in patients with opioid dependence. Polysubstance dependence was an exclusion criteria in one study (46,47), while cocaine use and HBV/HCV was reported for half of the sample at baseline in the other study (48,49). Standardized psychosocial counseling was provided in one study (46), while it was not reported by the other study (48). After 14-22 weeks of treatment, SROM was superior to methadone on almost all measures of mental health including depressive symptoms, anxiety symptoms, overall mental health, and different subscales of SCL-27 (Table 1). Assessment of heterogeneity was not feasible as there were insufficient data for meta-analysis on the overlapping outcomes between the two studies.

#### Methadone dose 0 -Methadone dose 20/50

One included study compared 3 different doses of oral methadone, i.e. 0mg, 20mg, and 50mg, in patients with intravenous opioid dependence, who did not have chronic medical illness or major psychiatric comorbidities (Table 1) (29). Counselling services were available to all participants. No significant difference was reported in the improvement of depressive symptoms among the 3 different dosing groups after 20 weeks of therapy.

#### Methadone-Buprenorphine

Four studies compared oral methadone with oral buprenorphine in patients with opioid dependence who had to have no other substance dependence in two studies (50,51), and had moderate to high prevalence of using other substances in the other two studies (52-55). In three studies, patients also had to have no history of disulfiram, antipsychotic, or anti-convulsant use, or history of major mental illness, including schizophrenia (50,52,54). During the 3-6months of therapy, there was no significant difference between methadone and buprenorphine in an array of mental health outcomes, while two studies reported superiority of methadone in improving depressive, obsessive compulsive, and phobic anxiety symptoms (50), as well as psychological problems in the past 30 days measured at baseline and follow-up visits (52). One study reported superiority of buprenorphine to methadone in improving depressive symptoms (Table 1) (51).

#### Buprenorphine-Waitlist/Placebo

Four studies compared oral buprenorphine, depot buprenorphine, and buprenorphine/naloxone with either placebo (2 studies) or waitlist (2 studies) (Table 1) in patients with opioid use disorder, who were explicitly substitution treatment-free in the 1-3 months prior to beginning the study. In one study (56), all patients were polysubstance dependent, while in the other 3 studies patients with other substance use disorders or major psychiatric comorbidities were excluded and prevalence of other substance use was reported at baseline (32,57,58). Weekly individual drug counselling appears to be the only psychosocial care provided in one of the studies (32). No significant difference was reported between buprenorphine and placebo, during the 12-24 weeks of treatment, in improving overall mental health, number of anxiety/depression episodes in the past month, or incidence of suicide ideation/attempt. Buprenorphine was superior to waitlist during the 12 weeks of treatment in all mental health outcomes, including but not limited to overall mental health, mental health quality of life, anxiety, and depressive symptoms (Table 1).

### Quantitative synthesis

#### Network Meta-analysis

Based on the availability of data, NMA was applied for depressive symptoms and overall mental health symptomatology outcomes. For depression, 7 RCTs were included. As represented by the thickness of the connecting lines in the geometry of the network in Table 3, there was 1 study for HDM-DAM, 3 studies for methadone-DAM, 2 studies for methadone-buprenorphine, and 1 study for buprenorphine-waitlist. As represented by the size of the nodes, methadone has the most number of participants followed by DAM, buprenorphine, HDM, and waitlist respectively. Both fixed and random effects models had an acceptable level of convergence based on trace and density plots, as well as Potential Scale Reduction Factor (PSRF) values equal to 1.00, but the model fit improved substantially from the fixed effects model (DIC=78.2) to the random effects model (DIC=14.2). Although effect estimates (Table 3) showed higher point estimate effect sizes for all medications compared with waitlist/placebo for reducing depressive symptoms, the effect sizes were statistically significant only for DAM and buprenorphine in the fixed-effects model, and not significant for any of the medications in the random effects model. For overall mental health, 10 RCTs were included. As represented by the thickness of the connecting lines in the geometry of the network in Table 3, there was 1 study for HDM-DAM, 1 study for SROM-methadone, 4 studies for methadone-DAM, 1 study for methadone-buprenorphine, and 4 studies for buprenorphine-waitlist/placebo. As represented by the size of the nodes in the geometry of the network in Table 4, buprenorphine has the most number of participants followed by methadone, DAM, SROM, waitlist/placebo, and HDM. Both fixed and random effects models had an acceptable level of convergence based on trace and density plots, as well as PSRF values equal to 1.00, and the model fit was comparable between the fixed effects model (DIC=20.9) and the random effects model (DIC=20.2). The effect estimates (Table 3) showed significant effects for all medications in improving overall mental health symptomatology compared to waitlist/placebo in the fixed effects model, and a significant effect for buprenorphine, diacetylmorphine, and methadone in the random effects model, where the highest point estimate effect size was for diacetylmorphine followed by methadone and buprenorphine. Finally, based on GRADE guidelines, for all pairwise comparisons between substitution medications and placebo/waitlist, confidence in effect estimates of depressive symptoms and overall mental health was either low or very low (Table 3).

#### Direct Pairwise Meta-analysis

In the direct pairwise comparisons, meta-analysis was applied for depression (Figure 2), overall mental health symptomatology (Figure 3), ASI psychiatric section (Figure 4), and mental health quality of life (Figure 5). For depression, there was a trend towards higher effect of diacetylmorphine compared with methadone based on the results of two studies with low level of between study variance (Tau^2^) and inconsistency (I^2^) (Figure 2b), and significantly higher effect of buprenorphine compared with waitlist based on the results of one study (Figure 2d). Comparison of buprenorphine and methadone using three studies with high level of between-study variance and inconsistency (Figure 2c), and DAM with HDM using one study (Figure 2a) did not show any significant difference. For overall mental health symptomatology, DAM was significantly more effective than methadone based on the results of four studies with zero variance and inconsistency (Figure 3b), and buprenorphine was significantly more effective than placebo/waitlist based on the results of three studies with a moderate level of between study variance but high inconsistency (Figure 3e). Comparisons of DAM with HDM (Figure 3a), buprenorphine with methadone (Figure 3d), and SROM with methadone (Figure 3c), with one study for each comparison, did not show any significant difference. For ASI psychiatric section, DAM was significantly more effective than methadone based on the results of four studies with low variance and moderate inconsistency (Figure 4b). Comparisons of DAM with HDM (Figure 4a), and buprenorphine with waitlist (Figure 4c), with one study for each comparison, did not show any significant difference. For quality of life mental health section, buprenorphine was significantly more effective than waitlist based on the results of one study (Figure 5c). Comparisons of DAM with methadone using two studies with zero variance and inconsistency (Figure 5a), and SROM with methadone using one study (Figure 5b) did not show any significant difference. Sensitivity analysis was not applicable as only 1 study had an overall low risk of bias.

## Discussion

From the 19 studies included in this review, 15 studies were used in the analyses out of which 14 had high overall risk of bias. Network meta-analysis showed that buprenorphine, DAM, and methadone were superior to waitlist/placebo in improving overall mental health symptomatology. No treatment was superior to placebo in improving depressive symptoms according to the network meta-analysis. Direct pairwise meta-analyses showed that overall mental health symptomatology improved more in DAM than methadone, and the same was true for psychiatric status. Depressive symptoms improved more in Buprenorphine than waitlist or placebo, and the same was true for overall mental health symptomatology as well as mental health quality of life. The results of all other direct pairwise comparisons were not significant.

Previous systematic reviews did not report comparative outcomes and instead focused on longitudinal changes in measures of mental health (11,12). Feelemyer et al. (12), only included cohort studies from specific countries while Fingleton et al. (11), included both cohorts and clinical trials from any country. Both studies reported improved mental health for most medications, but in many of the included studies, medication treatment was provided in conjunction with psychosocial interventions, which makes it hard to conclude that the effects seen were solely attributable to the medications. In our study, we were able to provide an improved picture of potential impact of opioid substitution treatments on mental health by crossing-out the effects of psychosocial interventions as much as possible. The importance of our findings is several folds; beside the robust literature published on superiority of drug use outcomes and retention in treatment for substitution programs compared with abstinence-based methods like detoxification, superiority of opioid agonists to placebo/waitlist on mental health outcomes further emphasizes opioid medication continuation/compliance as an essential part of the treatment approach. Beyond that, higher improvement of mental health during treatment with some opioid agonists over the others in this review, e.g. DAM compared to methadone, alongside their previously reported superior effectiveness on drug use outcomes, implies their potential added benefits for treatment of patients with more severe opioid use disorder or with higher comorbidities. This is in line with suggestions from previous original research (59). Last but not least, the current state of available evidence on mental health outcomes in RTCs of substitution treatment, implies the necessity of more attention to the psychiatric assessments of these patients, and integration of addiction psychiatry services to the current treatment systems as well as training of addiction physicians for assessment of psychiatric comorbidities (60).

Previous comprehensive reviews rated risk of bias as medium or low for some of the studies included in this review, using an older version of Cochrane risk of bias tool and focusing on retention in treatment and adverse events as the outcomes of interest (61). In this review, we assessed mental health as a subjective outcome using the extensively-revised and more comprehensive new version of Cochrane risk of bias tool, which explains lower ratings for risk of bias in our study compared to previous reviews. It would be inherently very hard to completely eliminate some of the sources of bias in future studies, such as blinding for comparison of medications with physiologically different effects, e.g. buprenorphine and methadone, because an experienced patient recognizes differential medication effects. Meanwhile some major considerations are well applicable to most of the future studies. These include but are not limited to planning and reporting analysis methods in advance, dealing with missing data properly and applying intention to treat analysis, clarity in reporting mental health outcomes, and documenting and reporting quantity and quality of ancillary services.

This review was subject to some limitations. First, many of the RCTs of opioid substitution treatment that would be otherwise eligible did not measure or report any mental health outcomes to be included in this review. In this regard, our findings should be interpreted with caution. Second, in most studies the exact quantity and quality of utilized psychosocial services were not reported. Meanwhile, we should note that all these trials were randomized, with baseline characteristics similar among the trial arms in each study, and any routine psychosocial service was similarly available to all participants. Third, except for one study, all the included studies had a high risk of bias. Fourth, we could not formally assess the publication bias due to insufficient number of studies for each pairwise comparison. Meanwhile, in the majority of the included studies, mental health outcomes were among the secondary outcomes, and consequently less prone to influence the publication status. Fifth, we did not investigate the dose-response relationship between opioid medications and mental health outcomes which needs to be further investigated at the individual patients-level data in future RCTs.

Overall, our findings show that mental health improves in substitution treatment with major opioid agonists independent of psychosocial services, and this improvement is greater in DAM substitution treatment than methadone. Future studies may further explore the role opioid dosage and treatment duration on mental health outcomes at the individual patients level data. Recommendations for future research mainly concern design and conduct of clinical trials of substitution treatment rather than further reviews.

## Data Availability

All the data-sets supporting the findings of this study are available from the corresponding author, EMZ, upon reasonable request.

## Author contributions

EMZ had full access to all of the data in the study and takes responsibility for the integrity of the data and the accuracy of the data analysis. EMZ contributed to the conceptualization, designed the study, coordinated the study, carried out the statistical analysis, provided the first draft, and acted as co-senior reviewer in cases of disagreement. KZ, KY, MM, and JW carried out screening, data extraction, and quality assessment. They also revised the manuscript for important intellectual content. AM contributed to the design, provided methodological guidance, and revised the manuscript for important intellectual content. PB and UV provided essential data for some of the studies included in this review. PB, UV, CS, KJ, SA, and MK provided consultation during the study, contributed to the interpretation of results and revised the manuscript for important intellectual content. MK conceptualized the study, contributed to the design, helped with coordination of the study, and acted as co-senior reviewer in cases of disagreement.

